# Non-isolated neural tube defects with comorbid malformations are responsive to population-level folic acid supplementation in Northern China

**DOI:** 10.1101/2022.07.12.22277037

**Authors:** Xiaoyu Che, Jufen Liu, Gabriel L. Galea, Yali Zhang, Nicholas D. E. Greene, Le Zhang, Lei Jin, Linlin Wang, Aiguo Ren, Zhiwen Li

**Affiliations:** Institute of Reproductive and Child Health / Key Laboratory of Reproductive Health, National Health Commission of the People’s Republic of China; Department of Epidemiology and Biostatistics, School of Public Health, Peking University, Beijing 100191, P. R. China; Developmental Biology and Cancer Department, UCL Great Ormond Street Institute of Child Health, University College London, London WC1N 1E, United Kingdom

**Keywords:** Neural tube defects (NTDs), congenital malformation, comorbidity, folic acid, primary prevention, Northern China

## Abstract

**Background:** Comorbid congenital malformation of multiple organs may indicate shared genetic/teratogenic causality. Folic acid supplementation reduces population-level prevalance of isolated neural tube defects (NTDs), but whether complex cases involving independent malformtions are also repsonsive is unknown. We aimed to describe the epidemiology of NTDs with comorbid malformations in a Chinese population and assess the impact of folic acid supplementation.

**Methods:** Data from five counties in northern China were obtained between 2002 to 2021 through a population-based birth defects surveillance system. All live births, stillbirths, and terminations because of NTDs at any gestational age were recorded. NTDs were classified as spina bifida, anaecephaly or encephalocele. Isolated NTDs included spina bifida cases with presumed secondary malformations (hydrocephalus, hip dislocation, talipes). Non-isolated NTDs were those with independent concomittant malformations.

**Results:** A total of 296,306 births and 2,031 cases of NTDs were recorded in 2002-2021. 4.8 % of NTDs (97/2031) had comorbid defects, which primarily affected the abdominal wall (25/97), musculoskeletal system (24/97), central nervous system (22/97), and face (15/97). The relative risk of cleft lip or/and palate, limb reduction defects, hip dislocation, gastroschisis, omphalocele, hydrocephalus and urogenital system defects was significantly greater in infants with NTDs than the general population. Population-level folic acid supplementation significantly reduced the prevalence of both isolated and non-isolated NTDs.

**Conclusion:** Epidemiologically, non-isolated NTDs follow similar trends as isolated cases and are responsive to primary prevention by folic acid supplementation. Various clinically-important congenital malformations are over-represented in individuals with NTDs, suggesting common etiology.

## Introduction

Neural tube defects (NTDs) are severe congenital malformations of the central nervous system (CNS) which affect approximately 1 in every 1,000 birthds globally^1^. Open NTDs such as spina bifida and anecephaly are caused by failure of neural tube closure, which normally completes by around 30 days of gestation in humans. Some forms of closed NTDs, such as encephalocele, occur after neural tube closure but may be genetically related to open NTDs^2^. While anecephaly is universally fatal, spina bifida and encephalocele are often amenable to surgical correction. Malformations in other organ systems are important causes of morbidity and mortality in individuals who survive with NTDs. Independent, comorbid malformations may reflect pleomorphic effects of genetic/teratogenic insults on morphogenesis of multiple organ systems.

Some malformations occur secondary to the NTD, such as Chiari II malformation, hydrocephalus, congenital hip dislocation and talipes in individuals who have spina bifida^3–5^. Hower, animal studies strongly corroborate the potential for pleiomorphic effects of gene mutations and teratogen exposure on multiple organ systems. For example, mutations in the planar cell polarity pathway can cause NTDs, abdominal wall closure defects and limb reduction defects in mice^6, 7^ In humans, genetic predispositions contribute to NTD risk, but their penetrance is complex and involves gene/environment interactions. The best known environmental factor is maternal folate status. This is clearly shown by clinical trials and by the dramatic reduction in NTD prevalence in northern China from 12 % in 2004 before folic acid supplementation, to 3.2 % in 2014 after supplementation^8^.

Although most epidemiological studies focused on isolated NTDs, those with comorbidities are also clinically important. In countries where fetal surgery for spina bifida is available, independent comorbid malformations are typically a surgical exclusion criterion^9^ When counselling parents whose pregnancy has been affected by a non-isolated NTD^10^, the evidence for a beneficial effect of folic acid is less robust than for isolated NTDs. A previous study in the USA found a progressive reduction in NTD rates between 1992 and 2009, but no significant change in non-isolated NTD rates over the same period^11^. In mice, folic acid does not rescue NTDs caused by mutations in some genes^12^ potentially changing the spectrum of associated malformations in supplemented populations.

Population differences might begin to explain variability in previous estimates of non-isolated NTD prevalence. The proportion of NTDs described as having comorbid malformations in community- or hospital-based studies ranges from 9.1% to 66%^13–19^. Our previous pathological anatomy study showed that 75.8% of NTD cases had additional malformations, of which the majority were musculoskeletal or visceral defects^20^. However, the definition of comorbid malformations is not standardised across studies, with those using sensitive methodologies such as fetal autopsy potentially being biased by analysing more severe terminated or non-viable cases. Additionally, spina bifida in particular is known to cause secondary malformations due to CSF leakage during gestation and musculoskeletal malformations due to deinnervation.

To our knowledge, few studies have described the population-level epidemiological status of non-isolated NTDs, excluding potentially secondary malformations. Therefore, we aimed to assess the epidemiological characteristics of comorbid malformations associated with NTDs in Northern China over a time period spanning introduction of folic acid supplementation.

## Study Design

### Birth Defects Surveillance

Five counties in Shanxi province, namely Pingding, Xiyang, Taigu, Zezhou, and Shouyang were included in the population-based birth defects surveillance system in the current study. The system was established in the early 2000s, and more than 20,000 births were recorded each year. All pregnant women who resided in the study area for more than 1 year were monitored. All live births (births of 28 or more complete gestational weeks), all stillbirths of at least 20 weeks’ gestational age, and pregnancy terminations at any gestational age following the prenatal diagnosis of NTDs were recorded^21^. NTD diagnosis was conducted by local specialists in maternal-fetal medicine and confirmed by pediatricians in Peking University. The study protocol was reviewed and approved by the Institutional Review Board of Peking University.

### Case Classification

The present study includes NTDs cases observed between 2002 and 2021. In addition to NTDs, there were 20 types of birth defects monitored, including congenital hydrocephalus, cleft palate and/or cleft lip, congenital ear defects, oesophageal atresia/stenosis, anorectal atresia/stenosis, hypospadias, hydronephrosis, club foot, polydactyly, syndactyly, limb reduction defects, omphalocele, gastroschisis, conjoined twins, Down syndrome, and congenital heart disease, among others. We also included other defects types recorded in the database, such as exposed viscera and cystic hygroma.

All NTD cases were classified as isolated or non-isolated. Congenital hydrocephalus, club foot and congenital dislocation of hip are considered to be secondary to spina bifida and were therefore included in the isolated NTDs cohort, but were considered non-isolated when they occurred with cranial NTDs. Additional defects were classified according to the organ system primarily affected: central nervous system (CNS) defects, craniofacial defects, gastrointestinal system defects, musculoskeletal system defects, abdominal wall defects, urogenital system defects, and others. Due to the possibility of a case having multiple major defects, subgroup categories are not mutually exclusive.

### Statistical Analysis

For NTDs, three types of prevalence were calculated: prevalence of total NTDs, isolated NTDs, and non-isolated NTDs. In calculating these indicators, the denominator remains the same: the total number of all live births. For other associated defects, two types of prevalence were calculated: prevalence of associated defects in the NTD group (the numerator is the number of cases with NTDs and associated defects; the denominator is the number of NTD cases) and prevalence of associated defects in the non-NTD group (the numerator is the number of cases with associated defects but no NTDs; the denominator is the number of live births without NTDs).

Cochran-Armitage Trend Tests were used to analyze the trend of prevalence of total, isolated and non-isolated NTDs. Chi-square tests were used to compare the effects of the residence of the mother, infant sex, gestational weeks and delivery time on isolated and non-isolated NTDs. Chi-square tests were also used to compare the prevalence of isolated and non-isolated NTDs before and after national folic acid supplementation. Poisson test was used to compare the prevalence of associated defects in NTDs group and non-NTDs group. The types and prevalence of defects associted with NTDs and the prevalence of three subtypes of NTDs combined with other defects were described. Two-tailed p<0.05 was considered statistically significant. All statistical analyses were performed using the R 4.0.2 software.

## Results

### The Prevalence, Types and Proportion of Defects Co-occurring with NTDs

From 2002 to 2021, a total of 296,306 births and 2,031 cases of NTDs were recorded in the Shanxi surveillance system. 28.4% (334/1175) of spina bifida cases suffered from defects believed to arise secondarily to their NTD, of which 293 had congenital hydrocephalus, 26 cases had clubfoot, and 15 cases had both congenital hydrocephalus and horseshoe foot. These “secondary” co-morbid malformations were included in the isolated NTD grouping.

Additionally, 4.8 % of NTDs (97/2031) had co-morbid defects not known to be secondary to spina bifida (Figure 1), including abdominal wall defects (25.8%, 25/97), musculoskeletal system defects (24.7%, 24/97), central nervous system defects (22.7%, 22/97), craniofacial defects (15.5%, 15/97), urogenital system defects (6.2%, 6/97), gastrointestinal system defects (2.1%, 2/97) and other defects (11.3%, 11/97). These 97 infants had 105 associated defects (some infants had defects in more than one site). There were six cases of spina bifida with two additional defects (limb reduction defect and gastroschisis; limb reduction defect and cleft palate; limb reduction defect and reproductive system defect; syndactyly and gastroschisis; cleft lip and palate and polydactyly; anorectal atresia or stenosis and reproductive system defect). There was also a case of multiple NTDs (anencephaly and spina bifida), with limb reduction defect and diaphragmatic hernia. Another case of multiple NTDs (anencephaly and encephalocele) had cleft lip and palate and clubfoot. In summary, co-morbid malformations in individuals who have NTDs are uncommon, but can affect multiple organ systems and have the potential to diminish quality of life of individuals who survive.

**Figure 1.**
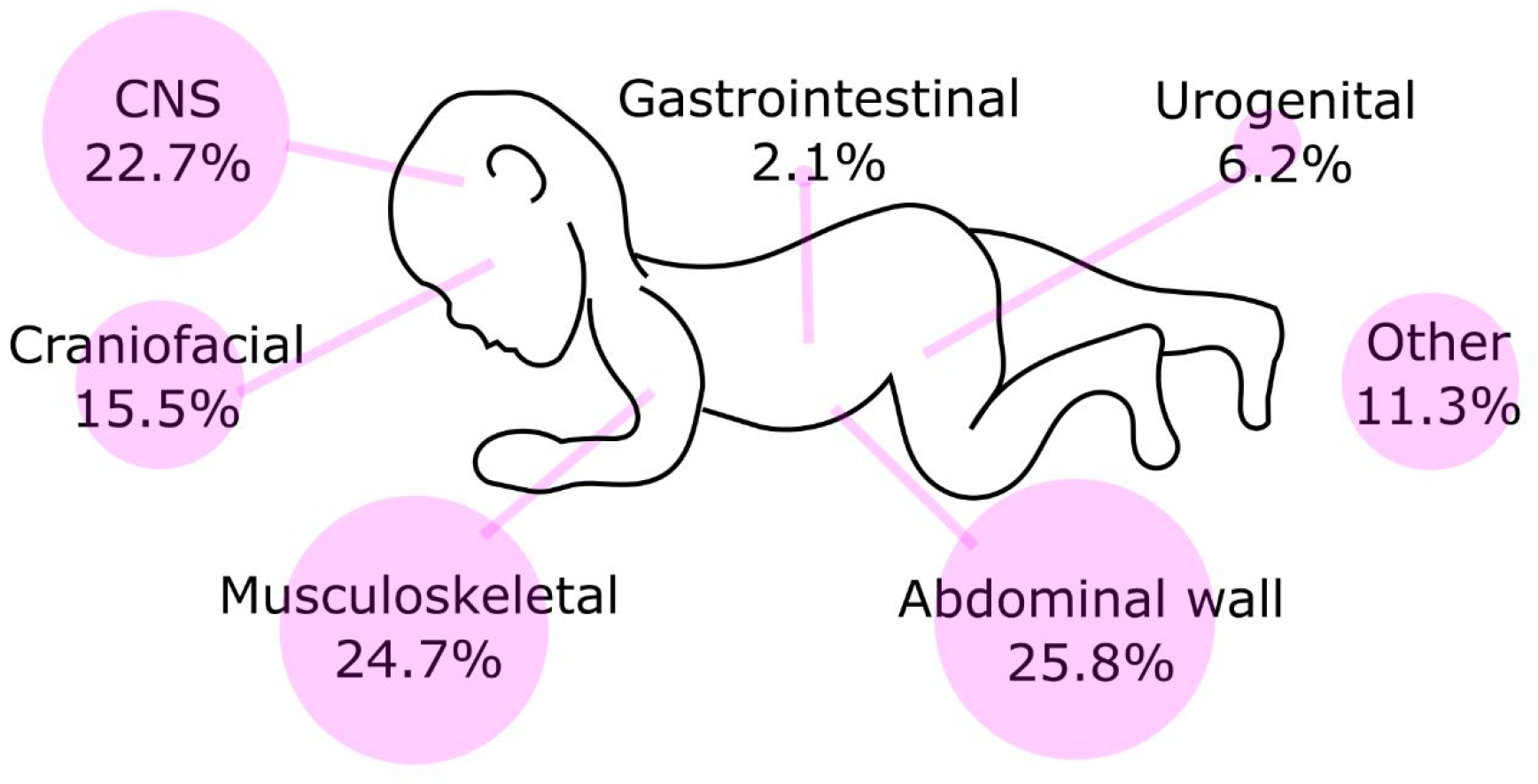
Associated defects among cases with NTDs in five counties in Shanxi province of northern China, 2002-2021

### Comorbid malformations over-represented in individuals with NTDs

We next assessed whether malformations of specific structures are over-represented in individuals who have NTDs, suggesting common genetic or environmental causation. The prevalence of cleft lip or/and palate (P<0.01), limb reduction defects (P<0.01), hip dislocation (P<0.05, excluding cases which also have spina bifida), omphalocele (P<0.01), gastroschisis (P<0.01), hydrocephalus (P<0.05, excluding cases which also have spina bifida) and urogenital system defects (P<0.01) is significantly higher in infants with NTDs than those born without NTDs (Figure 2).

**Figure 2.**
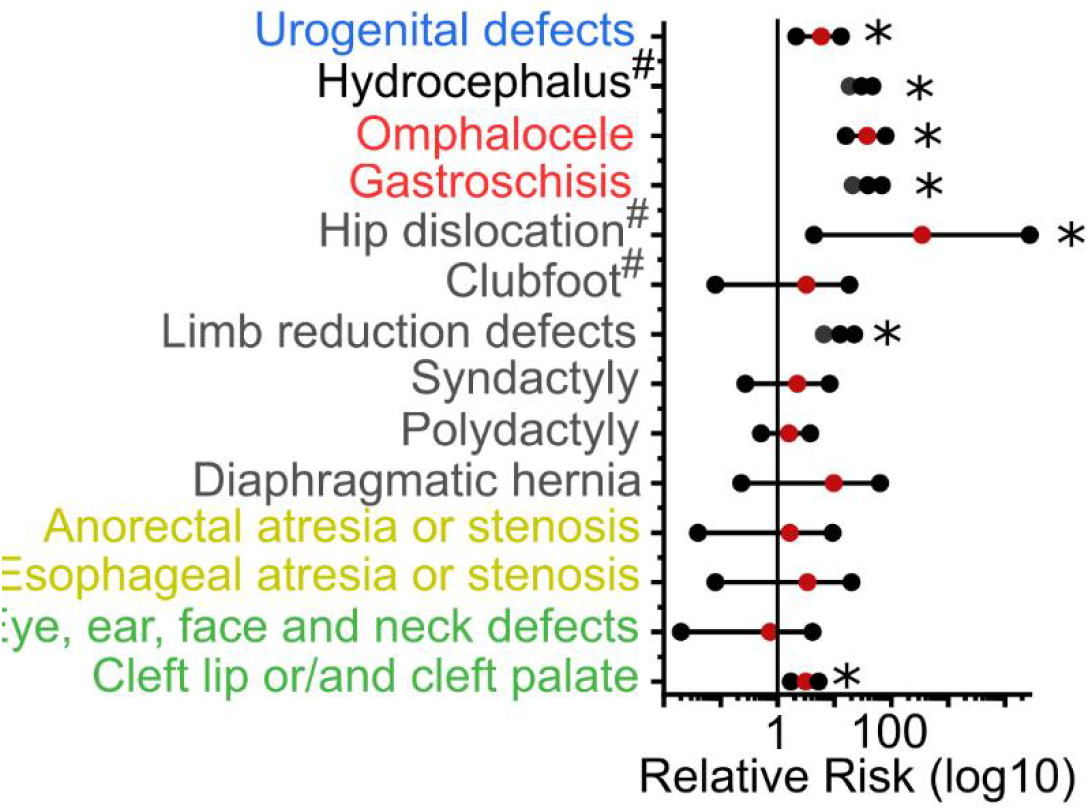
Relative risk of associated defects in NTDs group and non-NTDs group. # excludes cases with spina bifida

Associations with the three NTD sub-types were also tested individually (Table 1). Cleft lip and/or palate was more commonly observed in those with spina bifida (P < 0.01) or encephalocele (P < 0.05) than non-NTD controls. Limb reduction defects were more common in those with spina bifida (P < 0.01) or anencephaly (P < 0.01), whereas polydactyly was more common in those with encephalocele (P < 0.01). Diaphragmatic hernia was more common in those with anencephaly (P < 0.05). Hip dislocation was excluded from analysis in those with spina bifida but was over-represented in those with encephalocele (P < 0.01). Similarly, hydrocephalus was excluded from the analysis of those with spina bifida but was over-represented in those with sub-total anencephaly (P < 0.05) and encephalocele (P < 0.01). Omphalocele and gastroschisis were over-represented in those with either spina bifida (P < 0.01) or anencephaly (P < 0.01). Urogenital defects were specifically over-represented in those with spina bifida (P < 0.01).

**Table 1.**
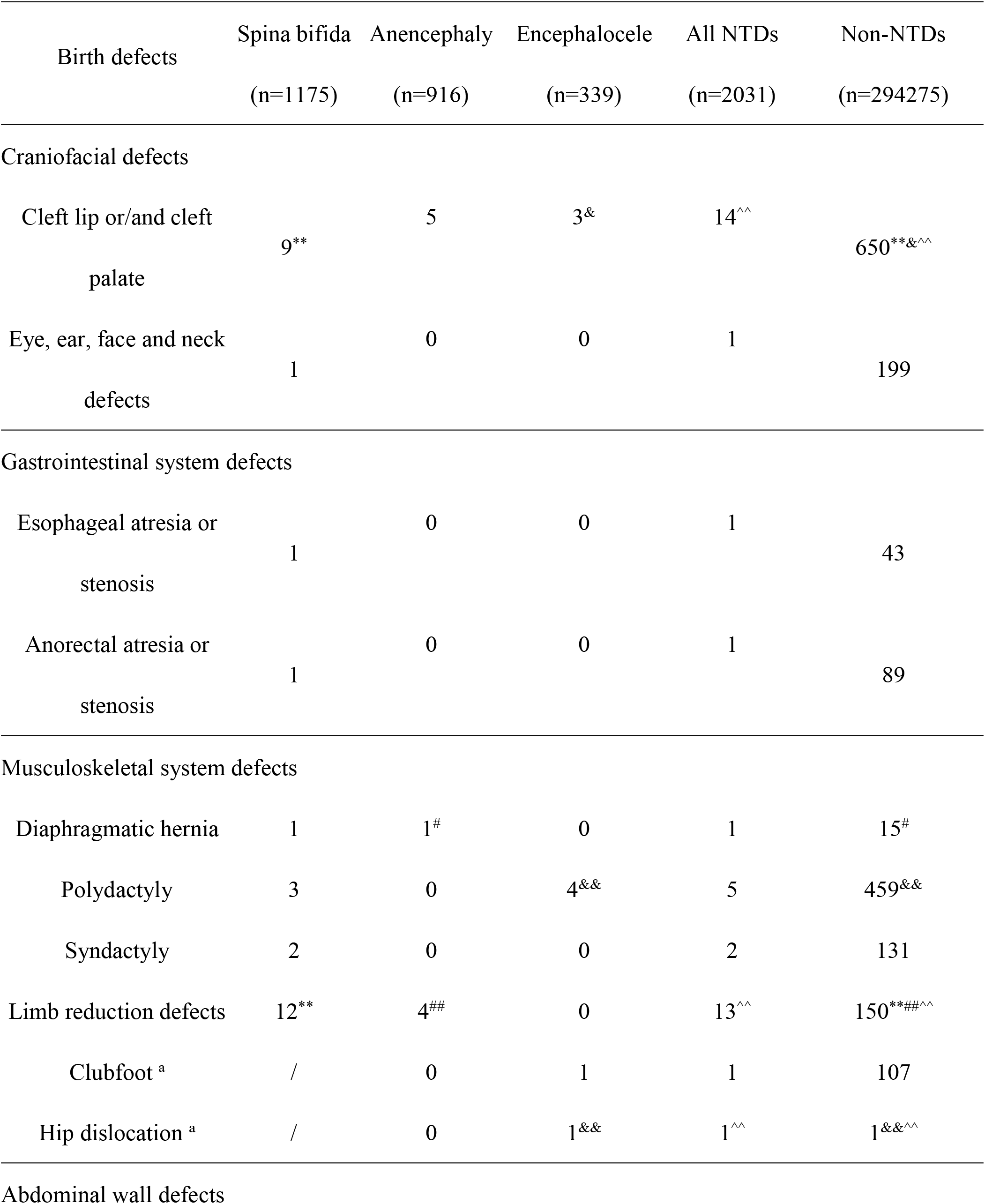
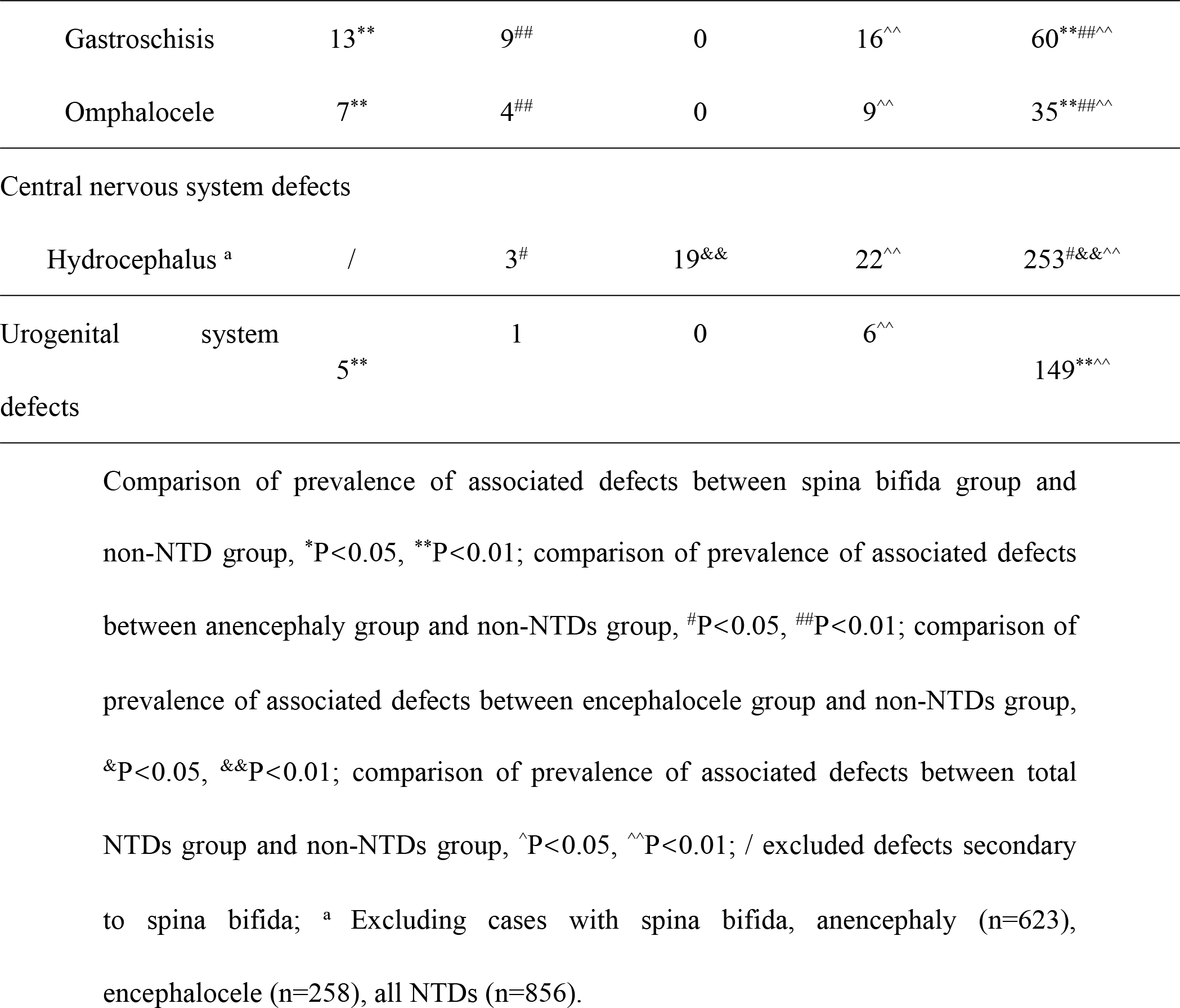
Associated defects in three NTDs by subtype groups and non-NTD group in five counties in Shanxi province of northern China, 2002-2021

### Epidemiological Characteristics of Isolated and Non-isolated NTD Cases

No differences in the distribution of maternal residence, infant sex and gestational weeks were observed between the isolated NTD group and those with NTDs and co-morbid malformations (Table 2). Individuals with encephalocele tended to be more likely to have additional malformations.

**Table 2.**
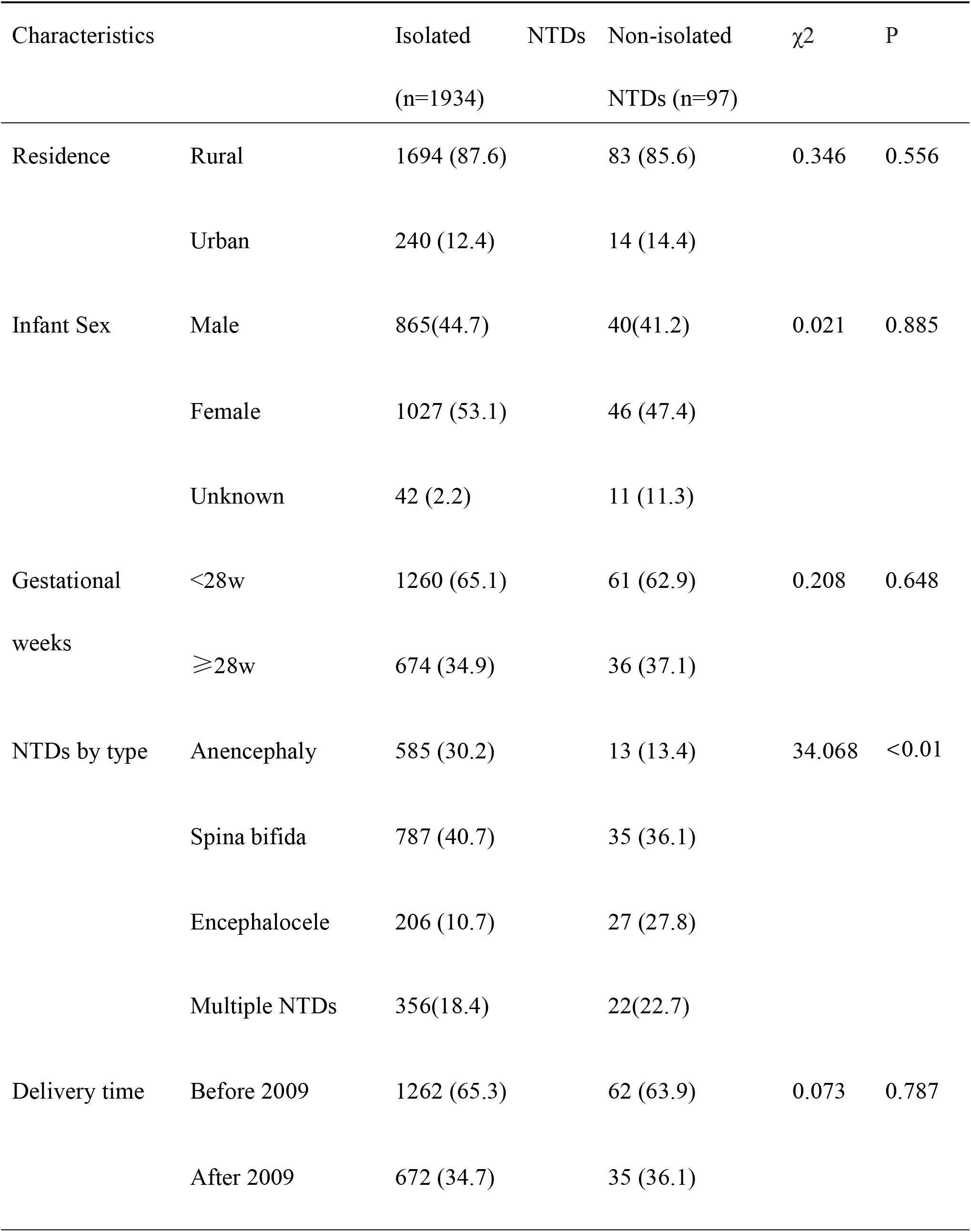
Characteristics of isolated and non-isolated NTD cases in five counties in Shanxi province of northern China, 2002-2021, N (%)

### Temporal Trends in the Prevalence of NTDs

We next asked whether non-isolated NTDs follow the same temporal change in prevalence as isolated NTDs which are the primary target of prevention programmes. The prevalence of total, isolated, non-isolated NTDs and multiple NTDs all decreased singificantly from 115.8/10,000, 109.7/10,000, 6.1/10,000 and 22.3/10,000 in 2003 to 15.5/10,000, 14.3/10,000, 1.2/10,000 and 2.4/10,000 in 2021 respectively (Figure 3A). The 3-year rolling average curves indicate progressively diminishing prevalence of both isolated and non-isolated NTDs (Figure 3B and 3C). Cochran-Armitage Trend Test results showed that the prevalence of total NTDs, isolated, non-isolated and multiple NTDs decreased significantly over the past two decades (P<0.01). Both isolated and non-isolated NTD subtypes all showed a significant downward trend, except for non-isolated spina bifida for which this did not reach significance (P>0.05)

**Figure 3.**
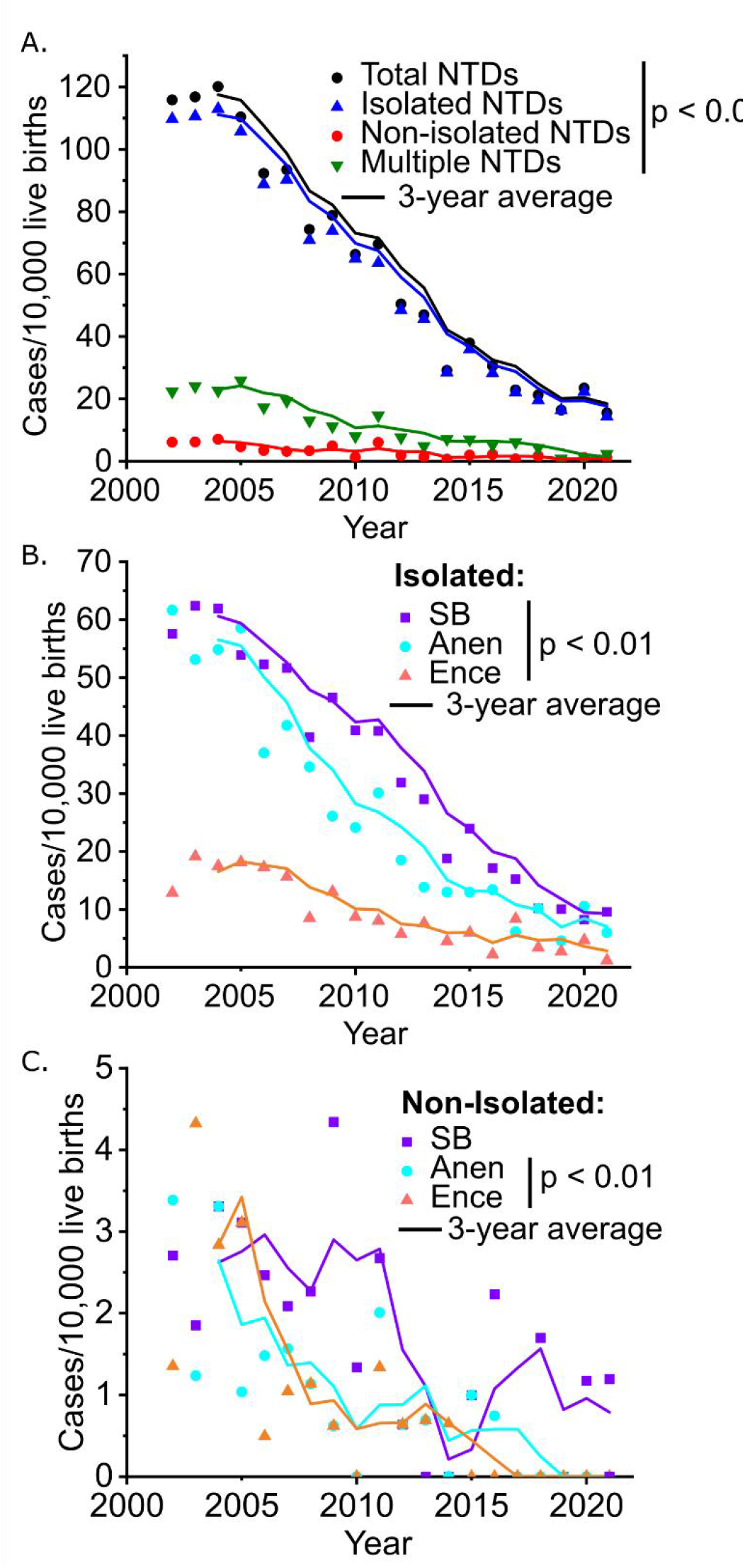
Time trends in the prevalence of NTDs in five counties in Shanxi province of northern China, 2002-2021 A. Total NTDs; B. Isolated NTDs; C. Non-isolated NTDs 2002, Zezhou, Pingding, and Taigu data are presented; 2003, Zezhou, Pingding, Taigu, and Shouyang data; other years, five counties.

### Prevalence of Isolated and Non-isolated NTDs before and after National Folic Acid Supplementation

The population survailance period inculdes various changes in public policy. The prevlance of non-isolated NTDs decreased from 5.14/10,000 live births in 2002–2008 (before population-level folic acid supplementation) to 2.85/10,000 in 2009–2015 (after population-level folic acid supplementation). After the implementation of the universal two-child policy, the prevalence of non-isolated NTDs dropped to 1.51/10,000 in 2016–2021 (Figure 4).

**Figure 4.**
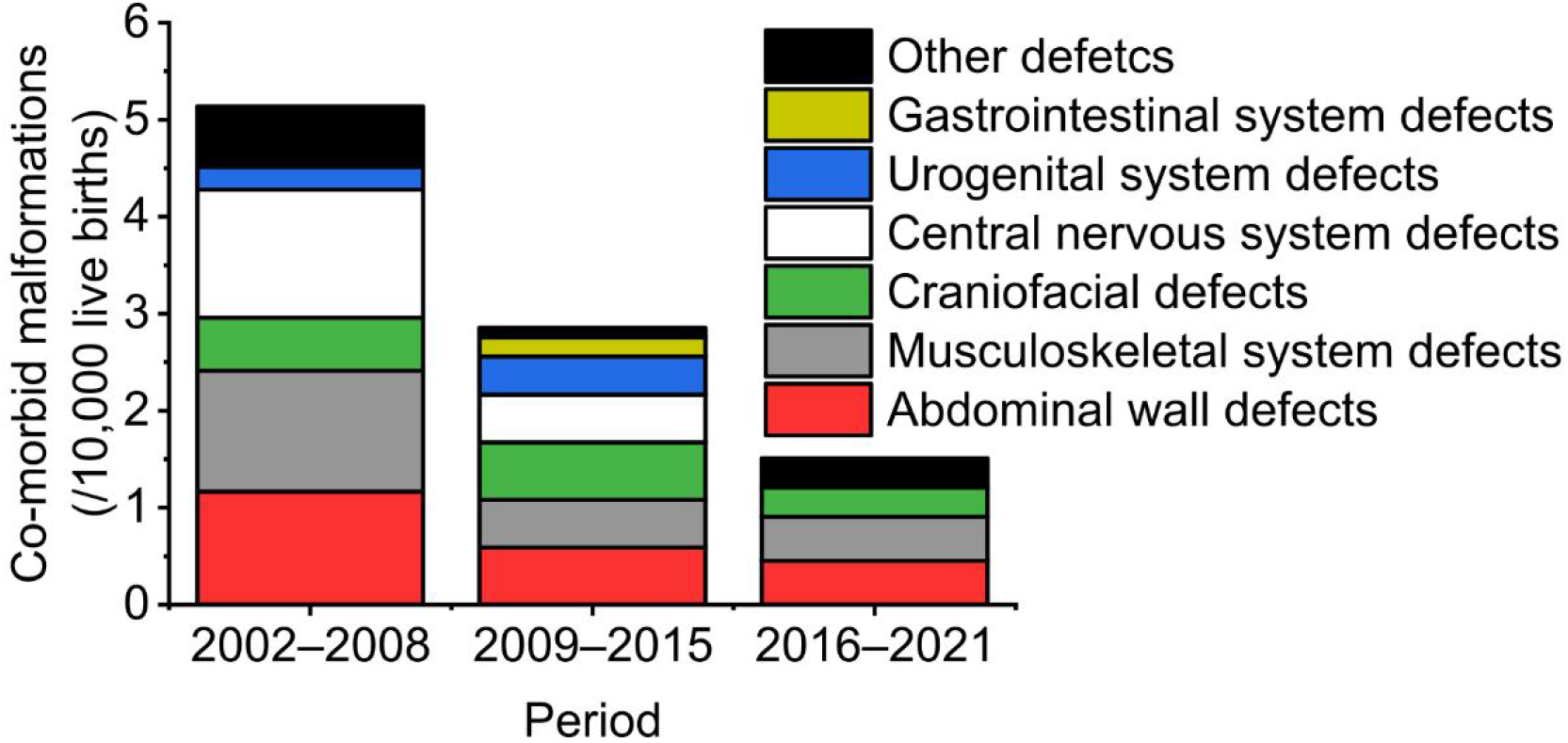
Prevalence of defects co-occurring with NTDs by period in five counties in Shanxi province of northern China, 2002-2021 The 3 periods were divided according to population policy and public strategy, i.e., 2002–2008 (before population-level folic acid supplementation), 2009–2015 (after population-level folic acid supplementation), 2016–2021 (population-level folic acid supplementation and universal two-child policy). On May 31, 2021, China officially implemented the universal three child policy.

After national folic acid supplementation, the prevalence of all NTD groupings, including both isolated cases and those with co-morbid malformations, decreased significantly (P<0.05) (Table 3). Non-isolated cases accounted for 4.7% of NTDs before folic acid supplementation and 4.9% of NTDs after supplementation.

**Table 3.** Prevalence of isolated and non-isolated NTDs before and after national folic acid supplementation in five counties in Shanxi province of northern China, 2002-2021, N (/10,000)

| Type of NTD | | Folic Acid Supplementation | | $\chi^2$ | P |
| --- | --- | --- | --- | --- | --- |
|  |  | Before (n=128477) | After (n=167829) |  |  |
| Total NTD | Isolated | 1262 (98.2) | 672 (40.0) | 379.954 | <0.01 |
|  | Non-isolated | 62 (4.8) | 35 (2.1) | 16.698 | <0.01 |
| Spina bifida | Isolated | 696 (54.2) | 423 (25.2) | 162.319 | <0.01 |
|  | Non-isolated | 33 (2.6) | 23 (1.4) | 5.528 | 0.019 |
| Anencephaly | Isolated | 633 (49.3) | 262 (15.6) | 273.760 | <0.01 |
|  | Non-isolated | 24 (1.9) | 8 (0.5) | 13.046 | <0.01 |
| Encephalocele | Isolated | 202 (15.7) | 105 (6.3) | 63.003 | <0.01 |
|  | Non-isolated | 26 (2.0) | 6 (0.4) | 18.709 | <0.01 |

## Discussion

NTDs remain prevalent and clinically important globally. Surgical advances have tremendously improved outcomes for individuals who have spina bifida, but the priority remains population-wide primary prevention. Cases which occur despite adequate maternal folate status are often assumed to reflect genetic/teratogenic causes not responsive to fortification or supplements. Using population-based surveillance data, this study reveals that 4.8% of NTDs have gross comorbid malformations, but that the epidemiology of non-isolated NTDs is comparable to isolated cases including a reduction in prevalence following population-wide folic acid supplementation.

Various authors have previously suggested that NTDs may be mechanistically linked to other malformations. Czeizel et al.^22^ hypothesized association with other defects of embryonic closure events, including cleft lip or/and cleft palate, posterior cleft palate, diaphragmatic hernia, and omphalocele, each being present in 1–6% of NTD cases. Opitz et al.^23^ suggested that midline structures share a particular common developmental vulnerability. Changes in the central nervous system may affect the lip and palate, diaphragm, heart, abdominal wall and genitalia that share the same midline. Although previous studies^15, 16^ provide no evidence to support the above two hypotheses, many of the conditions proposed are associated with NTDs in our study population. Compared with non-NTD controls, infants with NTDs were more likely to have cleft lip and/or palate, limb reduction defects, hip dislocation, gastroschisis, omphalocele, hydrocephalus and urogenital system defects.

These associations could be either genetic or teratogenic, if teratogen exposure spans the relevant susceptibility periods. For example, fetal valproate syndrome involves limb reduction defects as well as spina bifida. Genetic causes can also be shared between NTD subtypes and comorbid malformations: Grhl3^Cre^ deletion of Rac1 can cause abdominal call defects, spina bifida, exencephaly and/or encephalocele in mice ^24^. Encephalocele etiology is poorly understood and future work will be needed to explain our observation that they account for a larger proportion of non-isolated NTD cases.

Our results showed that there are no significant differences between isolated and non-isolated NTDs in key demographic characteristics, including residence of the mother, infant sex, subtype of NTDs, gestational weeks and delivery time, which suggested that non-isolated NTDs are more likely to be caused by multiple genetic factors. Due to the serious consequences of non-isolated NTDs, we need to further clarify the cause and identify other ways to reduce the risk of non-isolated NTDs. Further prospective cohort or case-control studies are needed to detect influences by more factors such as maternal diseases, childbirth history, family history and so on.

An unanswered question is whether folic acid prevents both NTDs and their comorbid malformations, or only the NTDs. Our study showed that the prevalence of all types of NTDs decreased significantly after national folic acid supplementation. A recent study in China also showed that periconceptional folic acid use prevents both rare and common NTDs^25^. In our previous study, we found that periconceptional folic acid supplementation can prevent congenital limb reduction defects in people with extremely low folate concentrations in northern China^26^. Some previous studies showed that perinatal use of multivitamin supplements effectively reduced the risk of multiple birth defects, even after excluding NTDs^11^. However, several studies showed that folic acid supplementation can reduce the risk of NTDs, but cannot reduce the risk of other birth defects^27^.

The prevalence of total NTDs, isolated NTDs and non-isolated NTDs in China showed a downward trend, which also reflected the remarkable results of preventing and controlling NTDs in the five counties of Shanxi Province in recent 20 years. However, when we conducted trend test according to different subtypes, the prevalence of non-isolated spina bifida did not show a downward trend. A previous study in South Carolina showed that the prevalence of total NTDs and isolated NTDs decreased significantly from 1992 to 2009, but the prevalence of non-isolated NTDs did not change significantly^28^. It is worth noting that since the three different subtypes of NTDs may be caused by different genetic factors, it is necessary to analyze the epidemiological trends according to different subtypes. Therefore, our results suggested that there may be etiological heterogeneity between non-isolated spina bifida and isolated spina bifida. A previous epidemiological study of NTDs also showed that isolated NTDs were more sensitive to environmental factors, whereas non-isolated NTDs were sensitive to both environmental and genetic factors^29^. In summary, the etiology of non-isolated NTDs needs to be confirmed by more population studies and laboratory studies.

Our current study found that changes in healthcare policy had an impact on the prevalence of non-isolated NTDs. China implemented the folic acid supplementation policy in 2009, and compared with 2002-2008, the prevalence of non-isolated NTDs decreased in 2009-2015. After the implementation of the universal two-child policy in 2016, the proportion of older and high-risk pregnant women may increase, resulting in an increased risk of NTDs^30, 31^. Other studies have found the opposite association, with high birth prevalence of spina bifida among younger mothers^32^. Our study showed that after the implementation of the universal two-child policy, the prevalence of non-isolated NTDs continued to decline. We speculate that there are two possible reasons. First, population-level folic acid supplementation can effectively prevent non-isolated NTDs, so the universal two-child policy had little impact on the prevalence of NTDs. Secondly, the impact of two-child policy on the overall population is not yet clear. We did not observe a rising trend of birth population after 2016 in the five counties of Shanxi Province. It is worth noting that the universal three-child policy was implemented in China in 2021, which means that further research on birth defects is needed to evaluate any impact on health outcomes in pregnancy.

Our study has several strengths. First, the surveillance is population-based and used quality control to ensure data quality. Our previous retrospective study showed that the surveillance covered 95.6% of births^33^. Second, previous to the present study, only a few studies have reported the epidemiological characteristics of isolated and non-isolated NTDs in China. We provided more detailed information about associated defects.

Despite the clear strengths of our study, some limitations should be acknowledged. First, the study included data from only five counties, and the results were not representative of other provinces or countries. Second, this study did not collect information on demographic and socio-economic factors, which prevented us from exploring the risk factors associated with isolated and non-isolated NTDs. Third, the monitoring system mainly reported external defects, so we may have missed some internal defects.

## Conclusion

Epidemiologically, non-isolated NTDs follow similar trends as isolated cases and are responsive to primary prevention by folic acid supplementation. Various clinically-important congenital malformations are over-represented in individuals with NTDs, suggesting common etiology.

## Data Availability

All data produced in the present study are available upon reasonable request to the authors

## Notes

Funding: This work was supported by the National Key Research and Development Program, Ministry of Science and Technology from P.R. China (Grant No. 2021YFC2701001), and the Natural Science Foundation of China (No.81973056). GLG acknowledges funding from the Wellcome Trust (211112/Z/18/Z) and the NIHR Great Ormond Street Institute of Child Health.

### Competing Interest Statement

The authors have declared no competing interest.

### Funding Statement

This work was supported by the National Key Research and Development Program, Ministry of Science and Technology from P.R. China (Grant No. 2021YFC2701001), and the Natural Science Foundation of China (No.81973056). GLG acknowledges funding from the Wellcome Trust (211112/Z/18/Z) and the NIHR Great Ormond Street Institute of Child Health.

### Author Declarations

The Institutional Review Board of Peking University gave ethical approval for this work

## Reference

[1] Zaganjor I, Sekkarie A, Tsang BL, Williams J, Razzaghi H, Mulinare J, et al. Describing the prevalence of neural tube defects worldwide: a systematic literature review. PloS one. 2016;11:e0151586.

[2] Rolo A, Galea GL, Savery D, Greene ND, Copp AJ. Novel mouse model of encephalocele: post-neurulation origin and relationship to open neural tube defects. Disease models mechanisms. 2019;12:dmm040683.

[3] Juranek J, Salman MS. Anomalous development of brain structure and function in spina bifida myelomeningocele. Developmental disabilities research reviews. 2010;16:23–30.

[4] Oliver E, Heuer G, Thom EA, Burrows PK, Didier R, DeBari S, et al. Myelomeningocele sac associated with worse lower-extremity neurological sequelae: evidence for prenatal neural stretch injury? Ultrasound in Obstetrics Gynecology. 2020;55:740–6.

[5] Viehweger E, Kläusler M, Loucheur N. Paralytic dislocation of the hip in children. Orthopaedics Traumatology: Surgery. 2021:103166.

[6] Murdoch JN, Damrau C, Paudyal A, Bogani D, Wells S, Greene ND, et al. Genetic interactions between planar cell polarity genes cause diverse neural tube defects in mice. Disease models mechanisms. 2014;7:1153–63.

[7] Wang B, Sinha T, Jiao K, Serra R, Wang J. Disruption of PCP signaling causes limb morphogenesis and skeletal defects and may underlie Robinow syndrome and brachydactyly type B. Human molecular genetics. 2011;20:271–85.

[8] Liu J, Zhang L, Li Z, Jin L, Zhang Y, Ye R, et al. Prevalence and trend of neural tube defects in five counties in Shanxi province of Northern China, 2000 to 2014. Birth Defects Research Part A: Clinical Molecular Teratology. 2016;106:267–74.

[9] Adzick NS, Thom EA, Spong CY, Brock III JW, Burrows PK, Johnson MP, et al. A randomized trial of prenatal versus postnatal repair of myelomeningocele. New England Journal of Medicine. 2011;364:993–1004.

[10] Ravindra VM, Aldave G, Weiner HL, Lee T, Belfort MA, Sanz-Cortes M, et al. Prenatal counseling for myelomeningocele in the era of fetal surgery: a shared decision-making approach. Journal of Neurosurgery: Pediatrics. 2020;25:640–7.

[11] Botto LD, Olney RS, Erickson JD. Vitamin supplements and the risk for congenital anomalies other than neural tube defects. American Journal of Medical Genetics Part C: Seminars in Medical Genetics: Wiley Online Library; 2004. p. 12–21.

[12] Marean A, Graf A, Zhang Y, Niswander L. Folic acid supplementation can adversely affect murine neural tube closure and embryonic survival. Human molecular genetics. 2011;20:3678–83.

[13] Barisic I, Boban L, Loane M, Garne E, Wellesley D, Calzolari E, et al. Meckel–Gruber Syndrome: a population-based study on prevalence, prenatal diagnosis, clinical features, and survival in Europe. European Journal of Human Genetics. 2015;23:746–52.

[14] Group EW. Prevalence of neural tube defects in 20 regions of Europe and the impact of prenatal diagnosis, 1980-1986. Journal of Epidemiology Community Health. 1991:52–8.

[15] Stevenson RE, Seaver LH, Collins JS, Dean JH. Neural tube defects and associated anomalies in South Carolina. Birth Defects Research Part A: Clinical Molecular Teratology. 2004;70:554–8.

[16] Stoll C, Dott B, Alembik Y, Roth MP. Associated malformations among infants with neural tube defects. American Journal of Medical Genetics Part A. 2011;155:565–8.

[17] McDonnell R, Johnson Z, Delaney V, Dack P. East Ireland 1980-1994: epidemiology of neural tube defects. Journal of Epidemiology Community Health. 1999;53:782–8.

[18] Källén B, Robert E, Harris J. Associated malformations in infants and fetuses with upper or lower neural tube defects. Teratology. 1998;57:56–63.

[19] Moradi B, Shakki Katouli F, Gity M, Kazemi MA, Shakiba M, Fattahi Masrour F. Neural tube defects: distribution and associated anomalies diagnosed by prenatal ultrasonography in Iranian fetuses. Journal of Obstetrics, Gynecology Cancer Research. 2017;2:1–8.

[20] Y W, L J, J L, Y Z, Z L, A R. Comparative analysis of pathological anatomy results and clinical reports of neural tube defects. Chinese Journal of reproductive health. 2015;26:207–10.

[21] Liu J, Wang L, Zhang Y, Zhang L, Jin L, Li Z, et al. Selected Structural Birth Defects—Shanxi Province, China, 2000-2019. China CDC Weekly. 2020;2:718.

[22] Czeizel A, Opitz JM. Schisis-association. American journal of medical genetics. 1981;10:25–35.

[23] Opitz JM, Gilbert EF. CNS anomalies and the midline as a “developmental field”. American Journal of Medical Genetics. 1982;12:443–55.

[24] Greene ND, Massa V, Copp AJ. Understanding the causes and prevention of neural tube defects: Insights from the splotch mouse model. Birth Defects Research Part A: Clinical Molecular Teratology. 2009;85:322–30.

[25] Zhou Y, Crider KS, Yeung LF, Rose CE, Li Z, Berry RJ, et al. Periconceptional folic acid use prevents both rare and common neural tube defects in China. Birth Defects Research. 2022;114:184–96.

[26] Liu J, Li Z, Ye R, Ren A, Liu J. Folic acid supplementation and risk for congenital limb reduction defects in China. International journal of epidemiology. 2019;48:2010–7.

[27] López-Camelo JS, Castilla EE, Orioli IM. Folic acid flour fortification: impact on the frequencies of 52 congenital anomaly types in three South American countries. American Journal of Medical Genetics Part A. 2010;152:2444–58.

[28] Collins JS, Atkinson KK, Dean JH, Best RG, Stevenson RE. Long term maintenance of neural tube defects prevention in a high prevalence state. The Journal of pediatrics. 2011;159:143–9. e2.

[29] Khoury MJ, Erickson JD, James LM. Etiologic heterogeneity of neural tube defects: clues from epidemiology. American Journal of Epidemiology. 1982;115:538–48.

[30] Liu J, Song L, Qiu J, Jing W, Wang L, Dai Y, et al. Reducing maternal mortality in China in the era of the two-child policy. BMJ global health. 2020;5:e002157.

[31] Berihu BA, Welderufael AL, Berhe Y, Magana T, Mulugeta A, Asfaw S, et al. Maternal risk factors associated with neural tube defects in Tigray regional state of Ethiopia. Brain Development. 2019;41:11–8.

[32] Liu S, Evans J, MacFarlane AJ, Ananth CV, Little J, Kramer MS, et al. Association of maternal risk factors with the recent rise of neural tube defects in Canada. Paediatric Perinatal Epidemiology. 2019;33:145–53.

[33] Li Z, Ren A, Zhang L, Ye R, Li S, Zheng J, et al. Extremely high prevalence of neural tube defects in a 4-county area in Shanxi Province, China. Birth Defects Research Part A: Clinical Molecular Teratology. 2006;76:237–40.

